# Evaluating the accuracy of *Salmonella* Typhi Hemolysin E and lipopolysaccharide IgA to discriminate enteric fever from other febrile illnesses in South Asia

**DOI:** 10.1101/2025.06.20.25329792

**Authors:** Jessica C. Seidman, Kristen Aiemjoy, Mehreen Adnan, Irum Fatima Dehraj, Junaid Iqbal, Khalid Iqbal, Seema Irfan, Nahidul Islam, Md. Shakiul Kabir, Nishan Katuwal, Noshi Maria, Muhammad Ashraf Memon, Sira Jam Munira, Shiva Ram Naga, Sailesh Pradhan, Anik Sarkar, Rajeev Shrestha, Sony Shrestha, Syed Muktadir Al Sium, Krista Vaidya, Douglas Ezra Morrison, Alice S. Carter, Senjuti Saha, Dipesh Tamrakar, Mohammad Tahir Yousafzai, Denise O. Garrett, Stephen P. Luby, Farah Naz Qamar, Samir Saha, Jason R. Andrews, Richelle C. Charles

**Author notes:** Contributed equally (Jessica C. Seidman, Kristen Aiemjoy).

## Abstract

Existing methods to identify patients infected with Salmonella enterica Typhi (S. Typhi) or Paratyphi are inadequately accurate, affordable, and efficient. We evaluated the discriminatory power of antibodies to S. Typhi hemolysin E (HlyE) and lipopolysaccharide (LPS) in Bangladesh, Nepal, and Pakistan. Plasma concentrations of anti-HlyE and LPS IgA were measured in blood culture-confirmed enteric fever cases and in febrile controls with laboratory-confirmed alternative etiology. Receiver operating characteristic analyses showed that combining anti-LPS and HlyE IgA distinguished enteric fever cases from other febrile illnesses with an area under the curve (AUC) of 0.93. Anti-LPS IgA alone performed nearly as well (AUC 0.92). In children under 5, the combination outperformed individual biomarkers (AUC 0.96 vs. 0.94 (HlyE), 0.93 (LPS)) and was most accurate in Bangladesh and Pakistan compared to Nepal. These findings support anti-HlyE and LPS IgA ELISA as an accurate method to identify enteric fever in endemic settings.

## Introduction

Enteric fever is a systemic infection caused by the gram-negative bacteria *Salmonella enterica* serovars Typhi (*S*. Typhi) and Paratyphi, which are transmitted through contaminated food and water. Enteric fever is, therefore, common in many settings with inadequate sanitation (1). Accurate diagnosis is challenged by non-specific clinical presentation and poor performance of commercially available diagnostics (2–4). Definitive identification of enteric fever requires isolation of *S*. Typhi or Paratyphi from blood or bone marrow culture, necessitating microbiology laboratory capacity and several days to obtain results. Although blood culture is highly specific, it has poor sensitivity, is expensive and slow, and not widely accessible for many in communities where the disease is most common (5). Conventional and more complex Polymerase Chain Reaction (PCR) assays to detect *S*. Typhi and Paratyphi also generally have low sensitivity, are expensive and not suited to most resource constrained settings (6). The other most widely used diagnostic assay is the Widal serum-agglutination test, which has poor accuracy in endemic settings (7). A recent evaluation by FIND of commercially available *S*. Typhi rapid diagnostics found that all failed to perform with both sensitivity and specificity above 90% (8).

Two antigens, the pore-forming cytotoxin Hemolysin E (HlyE) and *S*. Typhi lipopolysaccharide (LPS), have shown promise for serodiagnostic assays for enteric fever in previous studies, but prior studies have been limited by small sample size and narrow geographic scope (9–11). In a pediatric Nigerian cohort, LPS and HlyE-specific IgA provided good discrimination between *S*. Typhi cases and controls with other bacteremias; the receiver operator characteristic (ROC) area under the curve (AUC) for LPS-specific IgA was 0.90 and somewhat lower for HlyE-specific IgA (AUC 0.74) (12). A prior study by our group, including *S*. Typhi and Paratyphi A cases and other bacteremias in Nepal, found that the combination of antibodies to HlyE and LPS IgA was 90% sensitive and 92% specific (AUC 0.95) (9). More recent work using the same biomarkers in a lateral flow assay found that the dual antigen diagnostic had high accuracy in both Pakistan (AUC 0.93) and Bangladesh (AUC 0.97) (13,14). Further studies are needed to evaluate the robustness of these biomarkers across age strata, diverse study populations, over time and in comparison with other common etiologies of febrile illness. This study addresses these gaps by evaluating the performance of HlyE and LPS IgA biomarkers across three countries, multiple age groups, and a range of alternative confirmed febrile illnesses.

The multi-site SeroEpidemiology and Environmental Surveillance (SEES) Study measured longitudinal anti-HlyE and LPS antibody responses in blood culture-positive enteric fever cases and estimated enteric fever seroincidence rates in endemic communities (15). In this substudy, we evaluated the ability of anti-HlyE and LPS IgA antibody levels at clinical presentation to distinguish blood culture-confirmed enteric fever cases from prospectively recruited patients with confirmed alternative etiologies of acute febrile illness in Bangladesh, Nepal and Pakistan. We also evaluated how the dynamics of convalescent antibody responses could influence test performance in high burden settings.

## Materials and Methods

### Ethical review

This study was reviewed and approved by the Institutional Review Boards or Ethical Review Committees of collaborating institutions: Bangladesh Institute of Child Health (BICH-ERC-01/02/2019), Nepal Health Research Council for Dhulikhel Hospital (391/2018), Aga Khan University (2019-0410-4188), Pakistan National Bioethics Committee (4-87/NBC-341-Amend-revised/19/81), Stanford University (39557) and MassGeneral Brigham (2019P000152).

### Study participants and procedures

Individuals were recruited from hospitals and clinical laboratories participating in an enteric fever surveillance study in Dhaka, Bangladesh (Bangladesh Shishu Hospital & Institute), Kathmandu and Kavrepalanchowk, Nepal (Kathmandu Medical College and Teaching Hospital, Dhulikhel Hospital, Kathmandu University Hospital), and Karachi, Pakistan (Aga Khan University Hospital, Kharadar General Hospital) (16). Cases were eligible if they were blood culture-positive for *S*. Typhi or Paratyphi A, enrolled in the SEES Study and had a baseline plasma sample (15). Alternative etiology controls were eligible if they presented with 3 or more days of fever (outpatient) or any fever duration (inpatient), and had laboratory confirmation of another etiologic agent. Patients with multiple pathogens detected could be included as controls, unless they had a positive blood culture for *S*. Typhi or Paratyphi within 30 days of enrollment (blood cultures were not performed on all controls). Controls were excluded if they did not meet the diagnostic criteria for a confirmed alternative etiology (Appendix Table 1).

**Table 1.** Characteristics of laboratory-confirmed enteric fever cases and alternative etiology controls.

|  | Bangladesh |  | Nepal |  | Pakistan |  | All |  |  |
| --- | --- | --- | --- | --- | --- | --- | --- | --- | --- |
|  | Case<br>(N=411) | Control<br>(N=79) | Case<br>(N=155) | Control<br>(N=102) | Case<br>(N=84) | Control<br>(N=82) | Case<br>(N=650) | Control<br>(N=263) | p value |
| **Age** |  |  |  |  |  |  |  |  |  |
| Median (Q1, Q3) | 5.4 (3.3, 8.0) | 3.0 (1.0, 7.0) | 19.4 (14.9, 25.7) | 28.0 (19.5, 38.0) | 9.0 (4.0, 17.0) | 29.5 (17.0, 39.8) | 7.0 (4.0, 13.0) | 22.0 (4.5, 34.0) | <0.001 |
| **Age Category (y)**<br>**n (%)** |  |  |  |  |  |  |  |  |  |
| <5 | 180 (43.8) | 50 (63.3) | 4 (2.6) | 7 (6.9) | 24 (28.6) | 9 (11.0) | 208 (32.0) | 66 (25.1) | <0.001 |
| 5-15 | 230 (56.0) | 28 (35.4) | 39 (25.2) | 13 (12.7) | 36 (42.9) | 10 (12.2) | 305 (46.9) | 51 (19.4) |  |
| 16+ | 1 (0.2) | 1 (1.3) | 112 (72.3) | 82 (80.4) | 24 (28.6) | 63 (76.8) | 137 (21.1) | 146 (55.5) |  |
| **Gender** |  |  |  |  |  |  |  |  |  |
| Male n (%) | 214 (52.1) | 45 (57.0) | 95 (61.3) | 54 (52.9) | 48 (57.1) | 69 (84.1) | 357 (54.9) | 168 (63.9) | 0.0162 |
| **Days of fever at presentation** |  |  |  |  |  |  |  |  |  |
| Median (Q1, Q3) | 4.0 (3.0, 6.0) | 3.0 (2.0, 4.0) | 4.0 (3.0, 5.0) | 3.0 (3.0, 6.0) | 6.0 (4.8, 8.0) | 5.0 (4.0, 7.0) | 4.0 (3.0, 6.0) | 4.0 (3.0, 6.0) | 0.15 |
| **anti-HlyE IgA concentration** |  |  |  |  |  |  |  |  |  |
| Median (Q1, Q3) | 27.6 (10.4, 57.3) | 2.1 (0.8, 5.5) | 19.2 (5.9, 50.0) | 4.7 (2.6, 8.1) | 36.5 (15.9, 102.3) | 3.0 (1.5, 5.0) | 26.4 (9.5, 59.1) | 3.4 (1.6, 6.3) | <0.001 |
| **anti-LPS IgA concentration** |  |  |  |  |  |  |  |  |  |
| Median (Q1, Q3) | 82.3 (31.9, 158.9) | 2.3 (0.5, 5.6) | 106.1 (32.6, 223.8) | 4.5 (3.1, 9.8) | 162.6 (73.5, 321.0) | 6.6 (3.6, 10.9) | 98.0 (35.3, 190.2) | 4.5 (2.2, 9.4) | <0.001 |

p values estimated for the comparisons between all cases and controls

Written informed consent was obtained from all adult participants (18 years or older) and from a parent/guardian for children under 18 years; participants 15-17 years old also provided written assent. At enrollment, patient demographics and symptom history were captured using a structured questionnaire.

### HlyE and LPS IgA ELISAs

Plasma separated from whole blood by centrifugation was stored at −70°C until testing. To measure anti-HlyE and LPS IgA antibody levels, we used kinetic enzyme-linked immunosorbent assays (ELISA) as previously published (detailed in the Appendix Methods) (15).

### Statistical Analyses

The primary objective of this analysis was to evaluate the discriminatory power of anti-HlyE and LPS IgA — individually and in combination — to distinguish enteric fever cases from febrile controls with confirmed alternative etiologies in endemic settings. We limited inclusion to participants aged ≤50 years who reported ≤14 days of fever at presentation with ELISA results for both antigens and used t-tests and chi-squared tests to compare case and control characteristics. Using receiver operator characteristic (ROC) analysis, we evaluated how well antibodies to HlyE IgA and LPS IgA alone and in combination were able to distinguish between cases and controls in endemic settings. To evaluate the combined performance of the antigen pair, we used the fitted values from a logistic regression model with anti-HlyE IgA and LPS IgA ELISA values as predictors. Biomarker performance was assessed with areas under the curve (AUC) overall and in stratified analyses by site, age category, and *Salmonella* serovar. Sensitivity, specificity and associated 95% CIs were calculated by bootstrapping. In a sensitivity analysis, we stratified by duration of fever at clinical presentation and compared AUCs using Delong’s method (17).

To estimate how long after infection patients would continue to test positive using these biomarkers, we assessed the time point at which convalescent antibody concentrations declined below positivity thresholds defined post hoc by maximizing balanced accuracy. Cutpoints were identified for the pooled data; details on the calculation of single and joint antigen cutpoints are presented in the Appendix. We characterized post-infection longitudinal antibody dynamics using hierarchical two-phase within-host models (15,18). The model distinguished between an initial active infection period, with an exponential antibody rise and a non-exponential decay period post-pathogen elimination. Time was measured since self-reported fever onset. We used a Bayesian framework to jointly estimate model parameters and hyperparameters with Markov chain Monte Carlo sampling. We then evaluated when median longitudinal response fell below individual and combined antigen thresholds identified in the cutpoint analysis.

All analyses were performed using R version 4.3.3; ROC curves, optimal cutpoints and associated performance metrics and confidence intervals were calculated using the cutpointR package version 1.1.2. Fixed sensitivity and specificity and associated confidence intervals were estimated using the pROC package v1.18.5. Markov Chain Monte Carlo sampling was implemented using the JAGS version 4.3.2.

## Results

### Participant characteristics

From the SEES Study, 679 blood culture-positive *S*.Typhi and Paratyphi A cases had a blood sample collected at baseline and 650 were eligible for inclusion in this sub-study (Figure 1). In Bangladesh (N=411), the median age was 5.4 years (IQR: 4.7) and 48% were female; in Nepal (N=155) the median age was 19.4 years (IQR: 10.8) and 39% were female; and in Pakistan (N=84) the median age was 9.0 years (IQR: 13.0) and 43% were female (Table 1).

**Figure 1.**
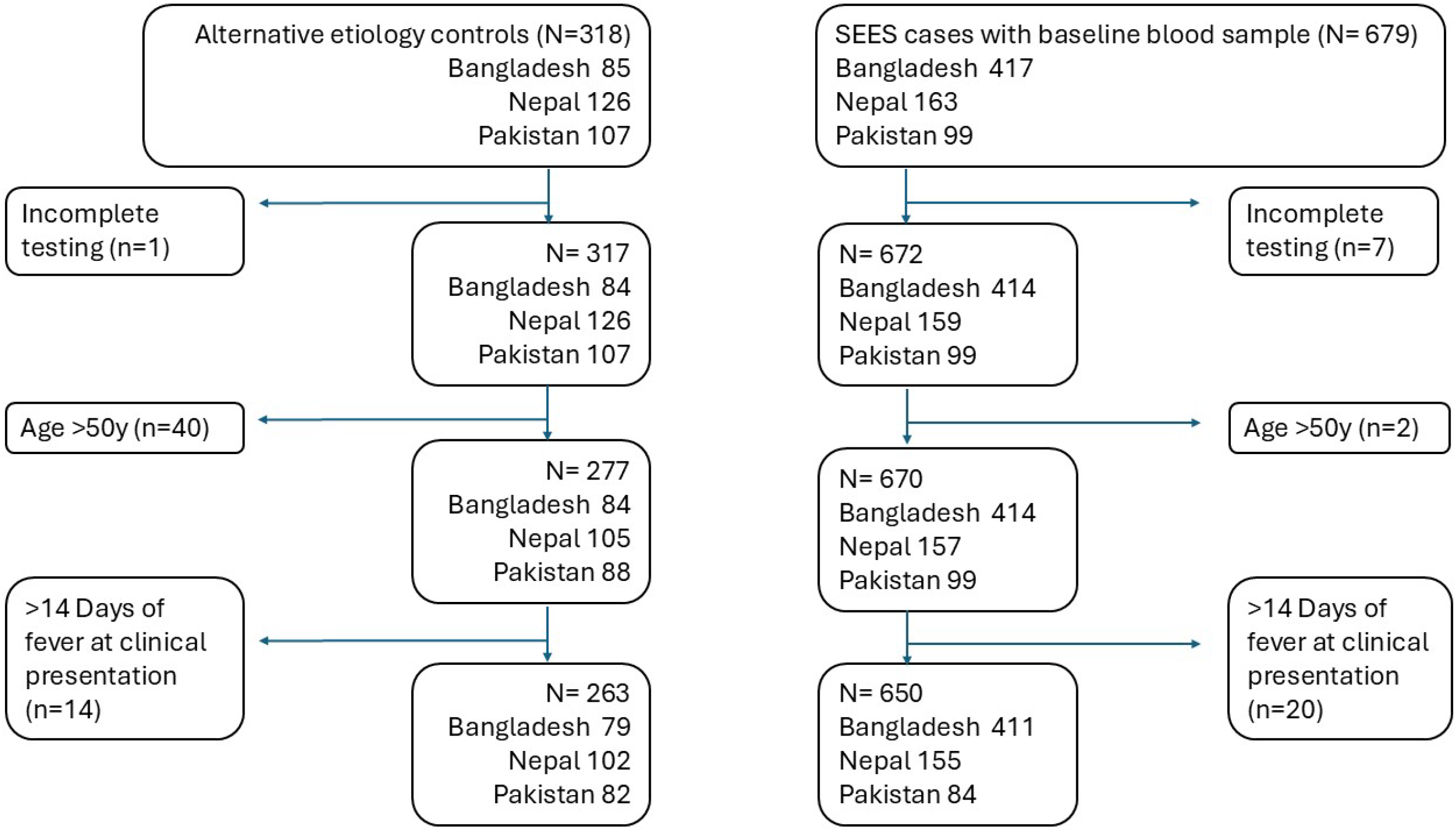
Participant enrollment flow

We recruited 318 febrile controls with laboratory-confirmed non-enteric fever etiology. After applying the age and fever inclusion criteria, 263 controls (79 from Bangladesh, 102 from Nepal, and 82 from Pakistan) were included in the analyses. The etiologies identified were dengue (n=128), SARS-CoV-2 (n=45), malaria (n=12), scrub typhus (*Orientia tsutsugamushi*, n=12), gram-negative (*Escherichia coli, Klebsiella pneumoniae, Acinetobacter* spp., *non-typhoidal Salmonella, Proteus mirabilis, Enterobacter cloacae;* n=40) and gram-positive bacteremia (*Staphylococcus aureus, Streptococcus* spp.; n=26). Across sites, the control population was significantly older than cases (median control age 22 years vs median case age 7 years; p<0.001).

We measured anti-HlyE and LPS IgA plasma antibody responses in febrile enteric fever cases and alternative etiology controls at presentation (Table 1, Figure 2). The median anti-HlyE and LPS IgA antibody levels were significantly higher in *S*. Typhi and Paratyphi A cases compared to controls (HlyE IgA: 26.4 (IQR 49.6) EU in cases vs 3.4 (IQR 4.7) EU in controls; LPS IgA: 98.0 (IQR 155.0) EU in cases vs 4.5 (IQR 7.2) EU in febrile controls (p<0.001)).

**Figure 2.**
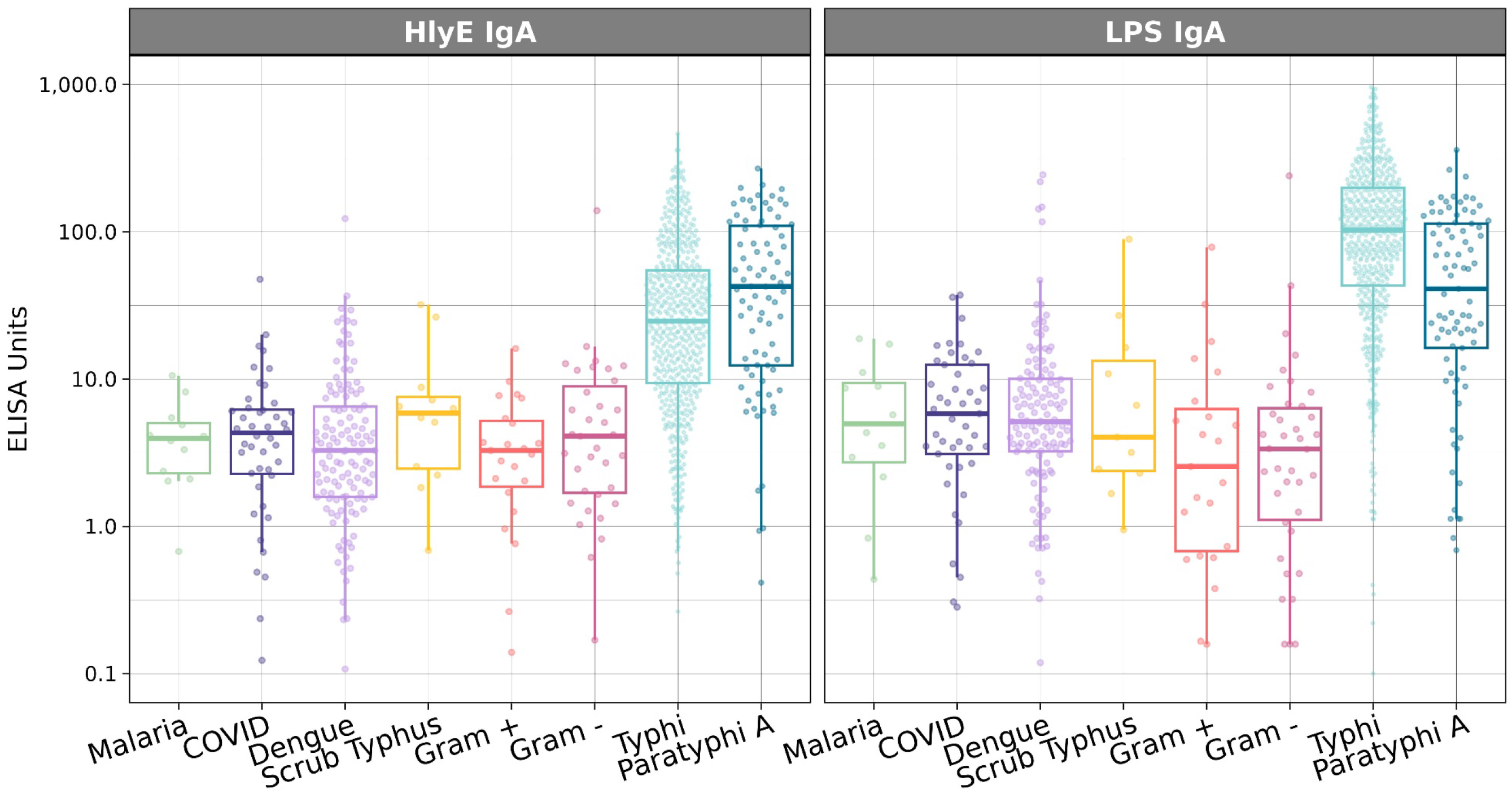
Distribution of anti-HlyE and LPS IgA antibodies among febrile cases by pathogen type. Boxplots show the distribution of plasma IgA responses against HlyE and LPS among *S*. Typhi and Paratyphi A cases and alternative etiology febrile controls.

### Discrimination of enteric fever cases using antibodies to HlyE and LPS IgA

We assessed the classification accuracy of anti-HlyE and LPS IgA overall and in stratified analyses by serovar, age and study site. In ROC analyses of the overall study population, the combined model including both HlyE and LPS IgA antibody concentrations yielded an AUC of 0.93 (95% CI: 0.91, 0.95) and at a sensitivity of 90% specificity was 86% (Figure 3, Appendix Table 2). The combined antigen model had slightly better performance than models using either antigen alone but anti-LPS IgA performed nearly as well as a solo biomarker (LPS AUC 0.92 (95% CI: 0.90, 0.94), HlyE AUC 0.87 (95% CI: 0.84, 0.89)). The combined antigen model was also slightly better at discriminating cases with Typhi (AUC 0.94 (95% CI: 0.92, 0.95)) vs Paratyphi A (AUC 0.90 (95% CI: 0.86, 0.95)) infections. However, as individual biomarkers, anti-HlyE IgA was marginally better at identifying paratyphoid cases than anti-LPS IgA (AUC 0.90 (95% CI: 0.85, 0.94) vs 0.84 (95% CI: 0.78, 0.90)).

**Figure 3.**
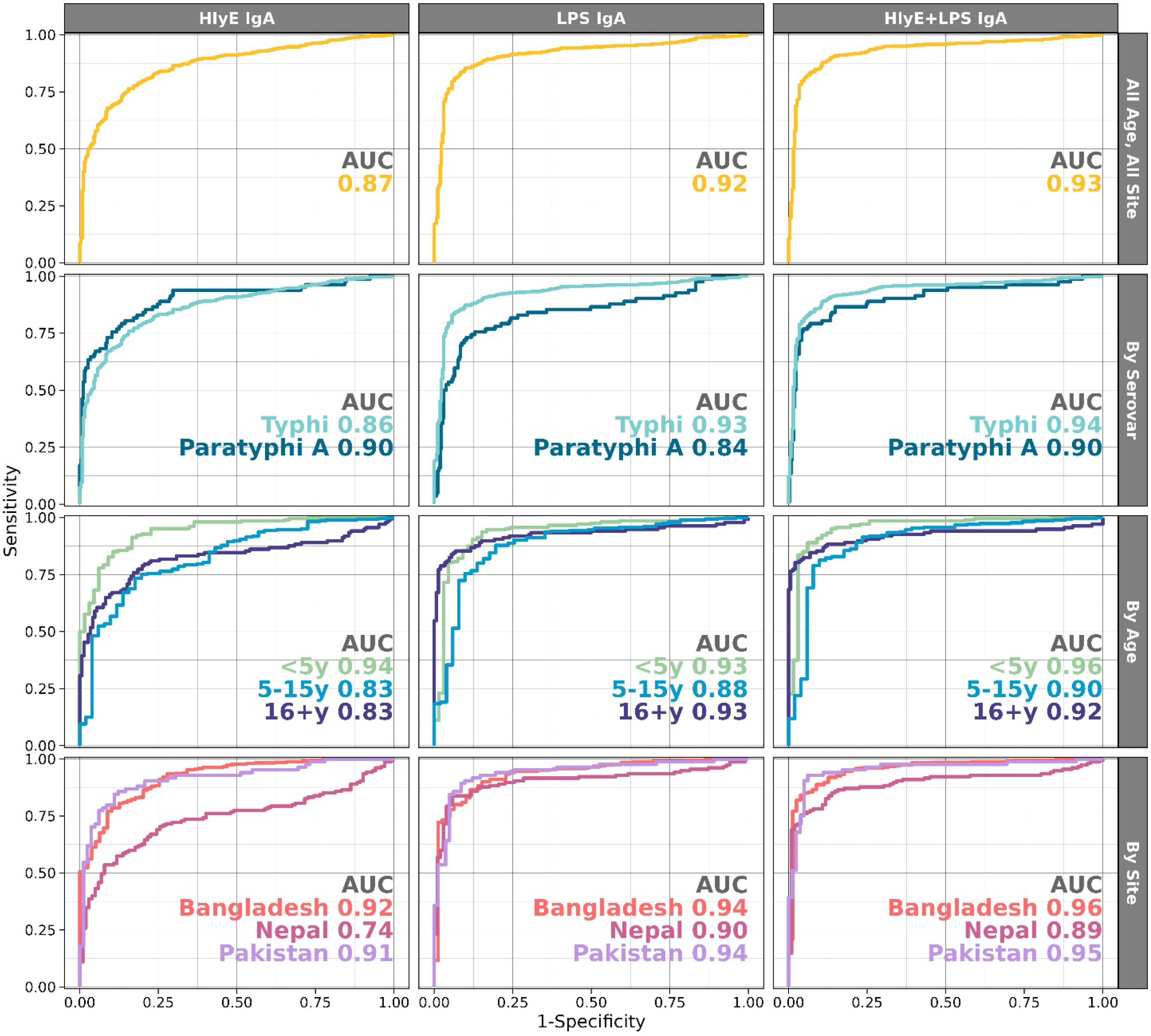
Diagnostic accuracy of IgA antibodies to *S*. Typhi HlyE and LPS alone and in combination was assessed by calculating the area under the receiver operating characteristic curve (AUC). Performance was assessed overall and in sub-analyses by *Salmonella* serovar, age category and study site.

We conducted stratified analyses to assay how well the biomarkers performed within site and age-specific strata. The biomarker pair worked best in the youngest children (age < 5y AUC 0.96 (95% CI: 0.92, 0.99)), a slight improvement over either antigen alone in this age group (HlyE: AUC 0.94 95%CI (0.91, 0.97); LPS AUC 0.93, 95%CI (0.90, 0.97)). The performance of the assay also differed by site; the Nepal cohort had the lowest AUC (0.89 (95% CI: 0.85, 0.94)) compared to Bangladesh (0.96 (95% CI: 0.93, 0.98)) and Pakistan (0.95 (95% CI: 0.92, 0.99)). In age and site-specific strata, the highest AUC was seen in Bangladesh in children <5 years (0.97) and the lowest in Nepal in children 5-15 years (0.82).

### Sensitivity analysis – duration of fever at clinical presentation

To investigate how the duration of fever reported at clinical presentation impacted accuracy, we stratified our dataset into fever tertiles (≤3, 4-5, 6-14 days of fever) and compared the performance of the biomarkers (Appendix Table 3). The combination of anti-HlyE and LPS IgA performed best at distinguishing cases from controls who presented with 4-5 days of fever (AUC 0.96, (95%CI 0.94, 0.98). In site-specific strata, there was no difference in accuracy by fever duration.

### Longitudinal antibody decay among convalescent cases

The modeled antibody dynamics revealed that anti-LPS IgA peaks earlier and decays more rapidly than anti-HlyE IgA (Figure 4, Appendix Table 4). This is reflected in its larger shape factor (r = 2.35 vs. r = 2.07) and slightly faster decay rate (alpha = 0.000355 /day vs. 0.000311 /day), indicating a more rapid initial decline in antibody concentration. Antibody responses to LPS IgA reached a significantly higher peak concentration (y1 = 232.6) at an earlier time point (t1 = 2.60 days), compared to the lower peak for HlyE IgA (y1 = 48.8) and later peak time (t1 = 3.59 days).

**Figure 4.**
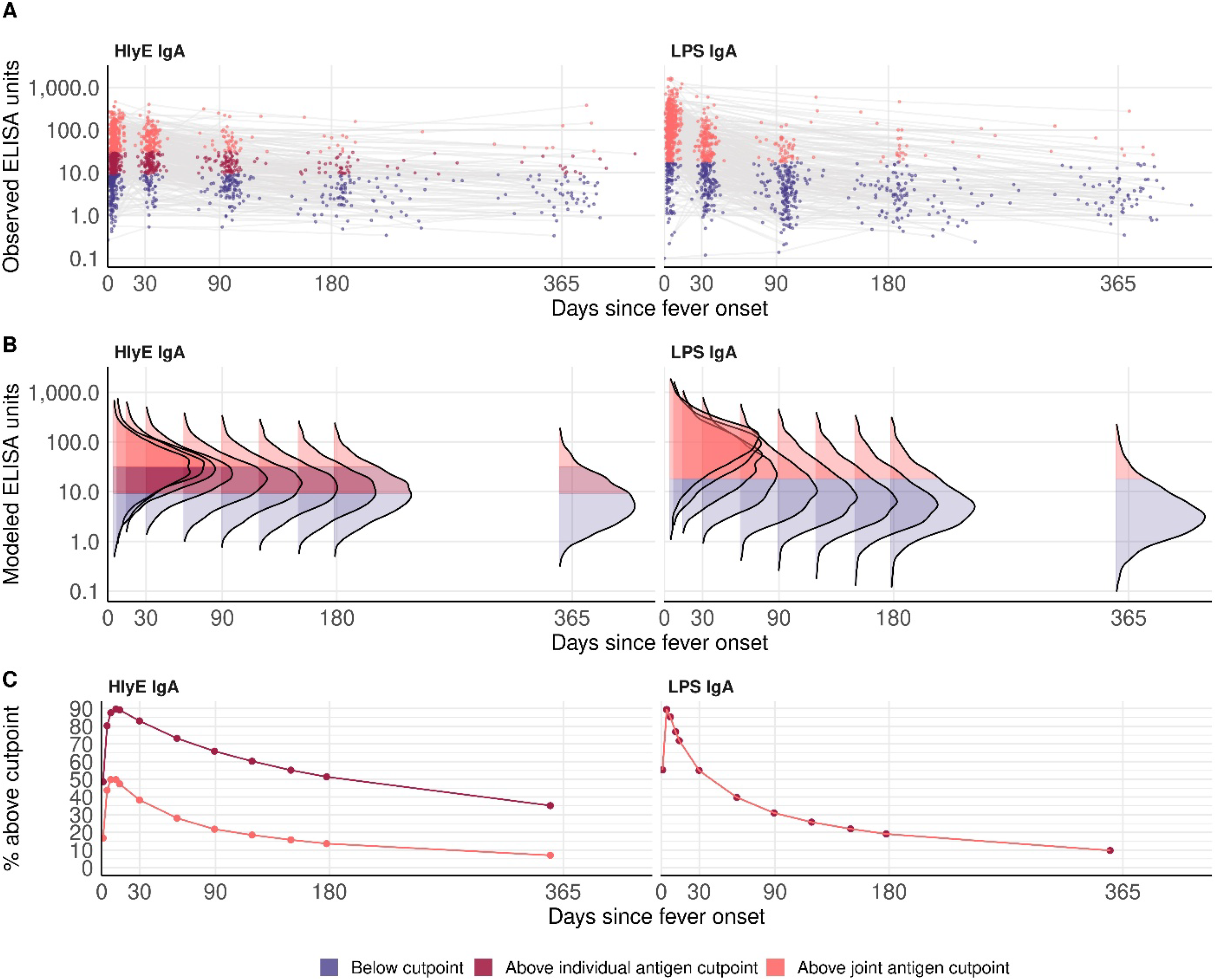
Antibody kinetics following infection: observed and modeled longitudinal anti-HlyE and LPS IgA antibody responses in enteric fever cases. A) Observed antibody responses from blood culture-confirmed enteric fever cases. B) Modeled distributions of antibody responses over one year following infection. C) The percent of modeled antibody responses above the individual and joint cutpoints for enteric fever case identification.

In order to estimate how long the antibody responses to *S*. Typhi or Paratyphi A infection remained above the case identification threshold, we compared the observed and modeled antibody decay curves to the antibody threshold values identified in the cutpoint analysis (Appendix Table 5, Appendix Figure 1). By 90 days after symptom onset, 78% and 69% of IgA antibody responses to HlyE and LPS respectively had fallen below the joint antigen cutoffs and thus would be classified as enteric fever negative by the combined biomarkers. The positivity threshold for anti-LPS IgA was the same when used alone or in combination with anti-HlyE IgA. For HlyE IgA alone, because the individual antigen threshold value was lower than the joint antigen threshold, antibody responses remained above the case identification cutoff for a longer duration. At 90 days following symptom onset, 66% of cases still had anti-HlyE IgA levels above the case identification threshold.

## Discussion

This study evaluated the performance of a serologic assay to detect enteric fever across three South Asian countries—Bangladesh, Nepal, and Pakistan—using well-characterized samples from blood culture-confirmed cases and febrile controls with laboratory-confirmed etiology. The assay, which measures IgA responses to *S*. Typhi LPS and HlyE, met the proposed minimum World Health Organization target product profile (TPP) thresholds for sensitivity and specificity (≥85%) (19). The performance of the assay varied by geographic setting and age group. These findings highlight the utility of context-specific validation when developing and deploying serologic tools for enteric fever diagnosis.

Differences in assay performance across sites may reflect underlying epidemiologic and immunologic factors, including the age distribution of cases, transmission intensity, and control group characteristics. Overall, the diagnostic accuracy was highest in Bangladesh and Pakistan with results in line with prior studies (9,12). However, the accuracy was lowest in Nepal, where enteric fever cases are older and the force of infection is lower. The performance of the assay differed from our prior study in Nepal in the same location (AUC 0.95) (9). It is possible that the Kathmandu valley has experienced changes in enteric fever transmission intensity since the prior study was performed.

The assay performed particularly well in children under five from Dhaka, Bangladesh (AUC 0.97), the region with the highest force of infection and clinical enteric fever incidence (16). In contrast, few samples from young children were collected in Nepal and samples of controls from Pakistan in this age group were limited. In both Pakistan and Nepal, assay performance was highest in the oldest age group (AUC 0.90 and 0.96, respectively).

We modeled the longitudinal persistence of anti-HlyE and LPS IgA to evaluate the duration of elevated antibody levels above cutpoints following acute infection. The higher peak responses and faster decay rate for anti-LPS IgA support its effectiveness as a diagnostic marker for recent infections, especially in settings with frequent *S*. Typhi exposure. In contrast, the slower decline and longer persistence of anti-HlyE IgA suggest its utility for seroepidemiological studies, offering insights into past exposures with a longer-lasting immune signature (15,20). Raising the anti-HlyE IgA cutoff value in the dual biomarker improved specificity by reducing false positives from longer-persisting antibody responses.

We have previously demonstrated that peak antibody responses to HlyE vary by age, which may explain the improved performance of LPS over HlyE, particularly at sites with higher forces of infection (i.e. Bangladesh and Pakistan compared to Nepal) (15). Including anti-HlyE IgA in the diagnostic provided only marginal benefit over anti-LPS IgA alone, as most cases with elevated antibodies to HlyE IgA also had a high concentration of anti-LPS antibodies. However anti-HlyE IgA was better than anti-LPS IgA at identifying Paratyphi A cases, which may be advantageous in contexts where Paratyphi A circulation is common. Our study was conducted in South Asia, where the prevalence of invasive non-typhoidal *Salmonella* (iNTS) infections is relatively low. In settings where the burden of iNTS disease is significant, particularly in Africa, including anti-HlyE IgA in a diagnostic panel may help improve the discriminatory power of the diagnostic since there is potential for cross-reactivity with LPS due to shared O-antigen. We have shown previously that HlyE IgA can better discriminate between these groups (15).

HlyE and LPS are not present in typhoid conjugate vaccine (TCV); thus, our selected biomarkers have diagnostic utility in areas where TCV has been introduced. However, these biomarkers are not able to distinguish *S*. Typhi from Paratyphi A infection, which has both clinical and public health implications. In our study sites, the circulating strains of Typhi and Paratyphi A have different patterns of resistance to antimicrobials which may impact treatment decisions by clinicians (21). TCV does not protect against paratyphoid infection; thus, it will be critical to understand the serovar-specific disease burden to inform introduction of the existing TCVs and forthcoming typhoid-paratyphoid combination vaccines, measure vaccine impact, and update treatment guidelines.

A limitation of our study is that although we enrolled a broad age range in both our case and control populations, we did not have equal representation in each age category at each site. Additionally, enteric fever cases were significantly younger than controls in the Nepal and Pakistan cohorts. This age difference may have reduced the apparent difference in biomarker levels between cases and controls. In selecting controls, we required laboratory confirmation of a non-enteric fever pathogen; however, blood culture was not performed for all controls and blood culture has low sensitivity (22). As a result, some controls may have been co-infected with *Salmonella*, which could have reduced our specificity estimates.

This study used a kinetic ELISA format assay which is best suited for well-provisioned laboratories. The end goal of typhoid diagnostic development is an accurate, rapid test that can be administered and interpreted with basic infrastructure and training. Kumar *et al* have developed a point-of-care test with these same antigens that can be used at lower-level clinical facilities with minimal laboratory set-up and training (23). This dual path, lateral flow assay demonstrated very high accuracy (AUC 0.98) when tested on adult samples from Bangladesh and Nepal and nearly as well in follow-up studies from Bangladeshi pediatric samples (AUC 0.97) and across a broad age range in Pakistan (AUC 0.92) (13,14).

Enteric fever continues to be a pervasive problem in many low-income settings, with an associated burden on patients, healthcare systems, and communities. There is an urgent need for better tools to identify enteric fever cases quickly, accurately and inexpensively. Furthermore, as the spread of antimicrobial-resistant *S*. Typhi limits the toolkit of effective therapies, the development of an improved diagnostic tool would lead to more appropriate treatment decisions and shorten the time to effective care. We have demonstrated that the combination of IgA antibodies to *S*. Typhi HlyE and LPS has excellent potential to identify febrile cases with enteric fever in endemic settings. Integration of this biomarker panel into point-of-care diagnostics or surveillance platforms could enhance case detection and guide treatment and prevention efforts.

## Data Availability

All data produced in the present study are available upon reasonable request to the authors

## Acknowledgments

We are extremely grateful to all the study participants who volunteered their time and specimens. This manuscript would not have been possible without the contributions of the large and dedicated laboratory, clinical and community research teams at the Child Health Research Foundation in Dhaka, Bangladesh, Dhulikhel Hospital and Kathmandu University Hospital in Dhulikhel Nepal, and Aga Khan University Hospital and Kharadar General Hospital in Karachi, Pakistan who worked tirelessly to conduct the SEES study. In particular we would like to thank the study teams from Bangladesh: Mohammad Saiful Islam Sajib, Nusrat Alam, Sultana Aflatun Rubana, Raktim Das, Farha Nusrat Zahan, Tanjila Akter, Sharif Husain, Khairun Naher, Kanis Fatema, Shamima Sultana, Masrufa Akhter, Jarin Sultana, Sathi Akter, Kristina Bain, Lima Akter, Shaswati Gain, Khursheda Afrin Khusi, Monalisa; Nepal: Sudan Maharjan, Lok Raj Bhatt, Natasha Shrestha, Shishir Ranjit, Anil Khanal, Bipin Khadka, Suman Shrestha, Pusp Raj Bhatt, Neeru Suwal, Suraj Jakibanjar; and Pakistan:. Yasmin Laddak, Kosar Riaz, Shazia Maqsood, Hira Asghar, Naik Banu, Afshan Piyar Ali, Hasina Wajid, Khalida Gul, Salima Shah, Samrina Karim, Faisal Hussain.

## Author contributions

JCS, KA, FNQ, DT, SaS, DOG, SPL, JRA, and RCC conceived and designed the study. MA, JI, SI, NI, NK, ASK, NM, SoS, and SMAS processed samples. IFD, KI, MSK, MAM, SJM, SRN, SP, RS, and KV collected samples and clinical metadata. JCS, KA, DEM, JRA, and RCC analyzed data. JCS, KA, ASC, IFD, MSK, SJM, SRN, KV, SeS, MTY, DT, DOG, SPL, FNQ, SaS, JRA, and RCC provided study oversight. JCS, KA and RCC prepared the figures and wrote the manuscript. All authors reviewed the manuscript and agree to its contents.

