## Appendix Methods for "Evaluating the accuracy of *Salmonella* Typhi Hemolysin E and lipopolysaccharide IgA to discriminate enteric fever from other febrile illnesses in South Asia"

Methods

*Detailed ELISA methods*

After coating plates with *S.* Typhi lipopolysaccharide (LPS) purified from strain Ty21a (2.5 µg/mL) or recombinant Hemolysin E (HlyE) (1 µg/mL), plasma samples were added in duplicate at dilutions of 1:1000 (LPS) or 1:500 (HlyE). Goat anti-human IgA conjugated to horseradish peroxidase (Jackson ImmunoResearch) was used to detect bound antibodies and peroxidase activity was measured at 450nm by using o-phenylenediamine. The maximum slope of the reaction over a three-minute period was extracted; results were reported as ELISA units (EU), the average of blank-adjusted sample values (milli-units per minute) divided by the mean value of the triplicate blank-adjusted positive controls and multiplied by 100.

*Cutpoint analysis*

We conducted a cutpoint analysis for each antigen individually and for the combination of both antigens. Individual cutpoints for each antigen were identified by finding the threshold that maximized the Youden's J statistic (sensitivity + specificity – 1). Separately, we then calculated sample percentiles of the ELISA values for each antigen and considered all 10,000 pairs of LPS and HlyE percentiles as cutpoints. For each pair of cutpoints, we classified samples as cases if the antibody concentration in the sample was greater than or equal to the corresponding cutpoint for *either* LPS IgA *or* HlyE IgA (or both). We then calculated the balanced accuracy ((sensitivity + specificity) / 2) for each pairwise combination of the percentiles and for the previously calculated individual antigen Youden's cutoffs. We identified the combinations of anti-HlyE and LPS IgA ELISA values that yielded the maximum balanced accuracy (rounded to two significant digits). When multiple combinations yielded the same balanced accuracy, we selected the threshold value that maximized sensitivity.
