## Appendix Table 1 for "Evaluating the accuracy of *Salmonella* Typhi Hemolysin E and lipopolysaccharide IgA to discriminate enteric fever from other febrile illnesses in South Asia"

**Appendix Table 1**. Diagnostic criteria and methods for alternative etiology febrile controls*

| Etiology | Diagnostic inclusion criteria |
| --- | --- |
| Dengue | Positive IgM serology OR NS1 antigen positive |
| Malaria | RDT positive OR blood smear positive |
| Scrub Typhus | Positive IgM serology |
| Other Bacteremia | Positive blood culture† excluding likely contaminants (e.g. coagulase negative *Staph spp*s, *Micrococcus*, *Bacillus sp*p, etc) |
| COVID-19 | Positive PCR or rapid antigen test |

* NS1= non-structural protein 1, PCR = polymerase chain reaction, RDT= rapid diagnostic test
†Blood culture was performed from whole blood using automated culture systems (BACTEC, Becton Dickinson, Franklin Lakes, NJ, USA; BacTAlert 3D, BioMérieux, Marcy-l'Étoile, France
