## Appendix Table 2 for "Evaluating the accuracy of *Salmonella* Typhi Hemolysin E and lipopolysaccharide IgA to discriminate enteric fever from other febrile illnesses in South Asia"

**Appendix Table 2**. Receiver Operating Characteristic analysis of IgA antibodies to HlyE and LPS overall and stratified by serovar, age category, and study site*

| Site / Age Category (y) | Serovar | N | | HlyE IgA | | | LPS IgA | | | HlyE + LPS IgA | | |
|---|---|---|---|---|---|---|---|---|---|---|---|---|
| | | **Case** | **Control** | **AUC (95% CI)** | **Sensitivity (95% CI) at 90% Specificity** | **Specificity (95% CI) at 90% Sensitivity** | **AUC (95% CI)** | **Sensitivity (95% CI) at 90% Specificity** | **Specificity (95% CI) at 90% Sensitivity** | **AUC (95% CI)** | **Sensitivity (95% CI) at 90% Specificity** | **Specificity (95% CI) at 90% Sensitivity** |
| All | Typhi + Paratyphi A | 650 | 263 | 0.87 (0.84, 0.89) | 0.68 (0.60, 0.75) | 0.58 (0.44, 0.69) | 0.92 (0.90, 0.94) | 0.85 (0.80, 0.89) | 0.80 (0.68, 0.87) | 0.93 (0.91, 0.95) | 0.86 (0.82, 0.91) | 0.86 (0.76, 0.91) |
| Overall | Typhi | 568 | 263 | 0.86 (0.84, 0.89) | 0.68 (0.59, 0.74) | 0.56 (0.42, 0.67) | 0.93 (0.91, 0.95) | 0.87 (0.83, 0.90) | 0.84 (0.77, 0.90) | 0.94 (0.92, 0.95) | 0.87 (0.83, 0.92) | 0.87 (0.79, 0.92) |
| | Paratyphi A | 82 | 263 | 0.90 (0.85, 0.94) | 0.74 (0.61, 0.84) | 0.73 (0.28, 0.84) | 0.84 (0.78, 0.90) | 0.72 (0.57, 0.82) | 0.37 (0.16, 0.75) | 0.90 (0.86, 0.95) | 0.80 (0.71, 0.88) | 0.70 (0.46, 0.89) |
| <5 | Typhi + Paratyphi A | 208 | 66 | 0.94 (0.91, 0.97) | 0.83 (0.64, 0.93) | 0.83 (0.73, 0.92) | 0.93 (0.90, 0.97) | 0.86 (0.74, 0.96) | 0.88 (0.77, 0.95) | 0.96 (0.92, 0.99) | 0.91 (0.83, 0.97) | 0.91 (0.82, 0.98) |
| 5-15 | | 305 | 51 | 0.83 (0.77, 0.89) | 0.58 (0.45, 0.76) | 0.49 (0.33, 0.65) | 0.88 (0.83, 0.94) | 0.75 (0.33, 0.89) | 0.73 (0.55, 0.86) | 0.90 (0.84, 0.95) | 0.81 (0.27, 0.89) | 0.76 (0.61, 0.88) |
| 16+ | | 137 | 146 | 0.83 (0.77, 0.88) | 0.66 (0.55, 0.74) | 0.18 (0.09, 0.60) | 0.93 (0.89, 0.96) | 0.86 (0.80, 0.92) | 0.82 (0.42, 0.94) | 0.92 (0.88, 0.96) | 0.86 (0.79, 0.92) | 0.76 (0.24, 0.94) |
| Bangladesh | | | | | | | | | | | | |
| Overall | Typhi + Paratyphi A | 411 | 79 | 0.92 (0.89, 0.95) | 0.76 (0.60, 0.84) | 0.75 (0.65, 0.84) | 0.94 (0.91, 0.96) | 0.82 (0.74, 0.91) | 0.84 (0.71, 0.92) | 0.96 (0.93, 0.98) | 0.88 (0.82, 0.94) | 0.89 (0.80, 0.95) |
| <5 | | 180 | 50 | 0.94 (0.90, 0.97) | 0.84 (0.72, 0.94) | 0.84 (0.70, 0.94) | 0.96 (0.93, 0.98) | 0.92 (0.79, 0.97) | 0.92 (0.80, 0.98) | 0.97 (0.95, 0.99) | 0.94 (0.86, 0.98) | 0.94 (0.84, 1.00) |
| 5-15 | | 230 | 28 | 0.88 (0.83, 0.94) | 0.64 (0.52, 0.83) | 0.61 (0.39, 0.79) | 0.89 (0.82, 0.96) | 0.76 (0.13, 0.87) | 0.68 (0.46, 0.86) | 0.92 (0.87, 0.98) | 0.83 (0.23, 0.92) | 0.79 (0.61, 0.93) |
| 16+ | | 1 | 1 | NA | NA | NA | NA | NA | NA | NA | NA | NA |
| Nepal | | | | | | | | | | | | |
| Overall | Typhi + Paratyphi A | 155 | 102 | 0.74 (0.68, 0.80) | 0.54 (0.41, 0.65) | 0.12 (0.05, 0.24) | 0.90 (0.86, 0.94) | 0.85 (0.77, 0.91) | 0.75 (0.23, 0.92) | 0.89 (0.85, 0.94) | 0.80 (0.72, 0.90) | 0.65 (0.15, 0.89) |
| <5 | | 4 | 7 | NA | NA | NA | NA | NA | NA | NA | NA | NA |
| 5-15 | | 39 | 13 | 0.62 (0.45, 0.79) | 0.31 (0.05, 0.59) | 0.15 (0.00, 0.46) | 0.83 (0.69, 0.97) | 0.54 (0.15, 0.95) | 0.38 (0.08, 0.92) | 0.82 (0.68, 0.96) | 0.64 (0.10, 0.90) | 0.38 (0.00, 0.85) |
| 16+ | | 112 | 82 | 0.78 (0.71, 0.84) | 0.59 (0.46, 0.70) | 0.07 (0.00, 0.39) | 0.92 (0.87, 0.96) | 0.84 (0.76, 0.92) | 0.78 (0.33, 0.93) | 0.90 (0.86, 0.95) | 0.83 (0.74, 0.91) | 0.67 (0.09, 0.91) |
| Pakistan | | | | | | | | | | | | |
| Overall | Typhi + Paratyphi A | 84 | 82 | 0.91 (0.87, 0.96) | 0.82 (0.68, 0.92) | 0.78 (0.39, 0.93) | 0.94 (0.90, 0.98) | 0.90 (0.77, 0.96) | 0.90 (0.70, 0.98) | 0.95 (0.92, 0.99) | 0.93 (0.85, 0.99) | 0.94 (0.80, 0.99) |
| <5 | | 24 | 9 | 0.94 (0.86, 1.00) | 0.75 (0.54, 1.00) | 0.78 (0.44, 1.00) | 0.77 (0.53, 1.00) | 0.12 (0.00, 0.83) | 0.56 (0.22, 0.89) | 0.83 (0.63, 1.00) | 0.29 (0.12, 0.96) | 0.78 (0.33, 1.00) |
| 5-15 | | 36 | 10 | 0.82 (0.65, 0.99) | 0.67 (0.08, 0.94) | 0.60 (0.10, 0.90) | 0.95 (0.88, 1.00) | 0.94 (0.58, 1.00) | 0.90 (0.60, 1.00) | 0.93 (0.83, 1.00) | 0.97 (0.39, 1.00) | 0.90 (0.60, 1.00) |
| 16+ | | 24 | 63 | 0.92 (0.83, 1.00) | 0.88 (0.67, 1.00) | 0.86 (0.14, 0.98) | 0.97 (0.92, 1.00) | 0.96 (0.83, 1.00) | 0.98 (0.40, 1.00) | 0.96 (0.90, 1.00) | 0.92 (0.79, 1.00) | 0.98 (0.21, 1.00) |

*AUC= Area Under the Curve, HlyE= Hemolysin E, LPS = Lipopolysaccharide, NA = Not applicable
