## Appendix Table 3 for "Evaluating the accuracy of *Salmonella* Typhi Hemolysin E and lipopolysaccharide IgA to discriminate enteric fever from other febrile illnesses in South Asia"

**Appendix Table 3**. Sensitivity analysis of the inclusion criteria for number of days of fever at clinical presentation

| | | | | HlyE IgA* | | LPS IgA | | HlyE + LPS IgA* | |
|---|---|---|---|---|---|---|---|---|---|
| **Site** | **Days of Fever** | **Case (N)** | **Control (N)** | **AUC** | **p value** | **AUC** | **p value** | **AUC** | **p value** |
| **All** | ≤3 | 200 | 123 | 0.82 (0.78, 0.87) | ref | 0.92 (0.89, 0.95) | ref | 0.93 (0.90, 0.96) | ref |
| | 4-5 | 223 | 69 | 0.89 (0.86, 0.93) | 0.02 | 0.94 (0.92, 0.97) | 0.22 | 0.96 (0.94, 0.98) | 0.06 |
| | 6-14 | 227 | 71 | 0.89 (0.84, 0.93) | 0.05 | 0.88 (0.84, 0.93) | 0.18 | 0.91 (0.87, 0.95) | 0.50 |
| **Bangladesh** | ≤3 | 120 | 47 | 0.87 (0.81, 0.93) | ref | 0.94 (0.89, 0.98) | ref | 0.94 (0.90, 0.99) | ref |
| | 4-5 | 146 | 19 | 0.96 (0.93, 1.00) | 0.01 | 0.96 (0.93, 0.99) | 0.43 | 0.98 (0.97, 1.00) | 0.09 |
| | 6-14 | 145 | 13 | 0.94 (0.89, 0.99) | 0.07 | 0.90 (0.84, 0.96) | 0.31 | 0.94 (0.90, 0.98) | 0.93 |
| **Nepal** | ≤3 | 70 | 56 | 0.71 (0.62, 0.80) | ref | 0.91 (0.85, 0.97) | ref | 0.89 (0.83, 0.95) | ref |
| | 4-5 | 48 | 19 | 0.74 (0.62, 0.85) | 0.70 | 0.92 (0.84, 0.99) | 0.91 | 0.92 (0.85, 0.99) | 0.56 |
| | 6-14 | 37 | 27 | 0.81 (0.70, 0.92) | 0.17 | 0.90 (0.81, 0.98) | 0.81 | 0.89 (0.79, 0.98) | 0.94 |
| **Pakistan** | ≤3 | 10 | 20 | 0.98 (0.95, 1.00) | ref | 0.98 (0.94, 1.00) | ref | 1.00 (1.00, 1.00) | ref |
| | 4-5 | 29 | 31 | 0.90 (0.82, 0.99) | 0.08 | 0.95 (0.89, 1.00) | 0.51 | 0.96 (0.89, 1.00) | 0.24 |
| | 6-14 | 45 | 31 | 0.90 (0.82, 0.97) | 0.89 | 0.90 (0.83, 0.97) | 0.29 | 0.92 (0.86, 0.99) | 0.44 |

* HlyE = Hemolysin E, LPS = Lipopolysaccharide, AUC = Area Under the Curve
