## Appendix Table 4 for "Evaluating the accuracy of *Salmonella* Typhi Hemolysin E and lipopolysaccharide IgA to discriminate enteric fever from other febrile illnesses in South Asia"

**Appendix Table 4**. Modeled longitudinal kinetic parameter estimates

| Parameter | Description | Units | HlyE IgA<br>Median (Q1, Q3) | LPS IgA*<br>Median, (Q1, Q3) |
|---|---|---|---|---|
| **Decay Rate (alpha)** | The rate at which antibody concentrations decline during the waning phase after the peak response.<br>-Smaller values indicate slower decay | Days | 0.00031<br>(0.00015, 0.00062) | 0.00035<br>(0.00011, 0.0010) |
| **Shape Factor (r)** | Describes the nonlinearity of antibody decay:<br>- When r>1r > 1, decay starts rapidly and slows over time, deviating from exponential decay<br>- Higher r values indicate faster early decay, transitioning to slower decay later | Dimensionless | 2.07<br>(1.80, 2.40) | 2.35<br>(2.01, 2.81) |
| **Time to Peak (t1)** | Represents the time taken to reach the maximum antibody concentration after symptom onset | Days | 3.59<br>(2.10, 5.94) | 2.60<br>(1.62, 4.14) |
| **Baseline Antibody Concentration (y0)** | Initial antibody concentration before infection | ELISA Units | 4.34<br>(1.98, 10.49) | 4.64<br>(2.44, 10.44) |
| **Peak Antibody Concentration (y1)** | Maximum antibody concentration achieved at peak | ELISA Units | 48.80<br>(25.61, 94.13) | 232.55<br>(125.95, 453.51) |

* HlyE = Hemolysin E, LPS = Lipopolysaccharide
