## Appendix Table 5 for "Evaluating the accuracy of *Salmonella* Typhi Hemolysin E and lipopolysaccharide IgA to discriminate enteric fever from other febrile illnesses in South Asia"

**Appendix Table 5**. Performance of Individual and Joint Antigen Cutpoints

| Performance measure | Individual Antigen | | Joint Antigen | |
|---|---|---|---|---|
| | HlyE IgA | LPS IgA | HlyE IgA | LPS IgA* |
| **Cutoff (EU)** | 9.23 | 18.05 | 31.21 | 18.05 |
| **Sensitivity** | 0.76 (0.65, 0.85) | 0.85 (0.78, 0.89) | 0.88 (0.85, 0.92) | |
| **Specificity** | 0.84 (0.71, 0.92) | 0.90 (0.81, 0.96) | 0.89 (0.84, 0.94) | |
| **Balanced Accuracy** | 0.80 | 0.88 | 0.89 | |

* HlyE = Hemolysin E, LPS = Lipopolysaccharide
