## Appendix Figure 1 for "Evaluating the accuracy of *Salmonella* Typhi Hemolysin E and lipopolysaccharide IgA to discriminate enteric fever from other febrile illnesses in South Asia"

**Appendix Figure 1**. Cutpoint analysis. Plots A, B, and C show ELISA values for IgA antibodies to HlyE and LPS from Typhi and Paratyphi A cases and alternative etiology controls. Case identification thresholds are indicated by overlaid lines and values classified as cases are shown in the shaded gray area. Individual antigen cutpoints selected by maximizing Youden's criteria are shown in A ( HlyE) and B (LPS). For the joint antigen cutpoint, sample generated percentiles of the ELISA values were calculated for each antigen and all 10,000 pairs of LPS and HlyE percentiles were considered as joint cutpoints. For each pair of cutpoints, samples were classified as cases if the antibody concentration in the sample was greater than or equal to the corresponding cutpoint for *either* anti-LPS IgA *or* HlyE IgA (or both). The balanced accuracy ((sensitivity + specificity) / 2) was calculated for each pairwise combination of the percentiles. In C, each cutpoint pair is shown as a circle on the larger plot with color indicating the balanced accuracy. The inset plot shows the optimal joint cutpoint and the classification rules can be visualized as rectangular classification boundaries.

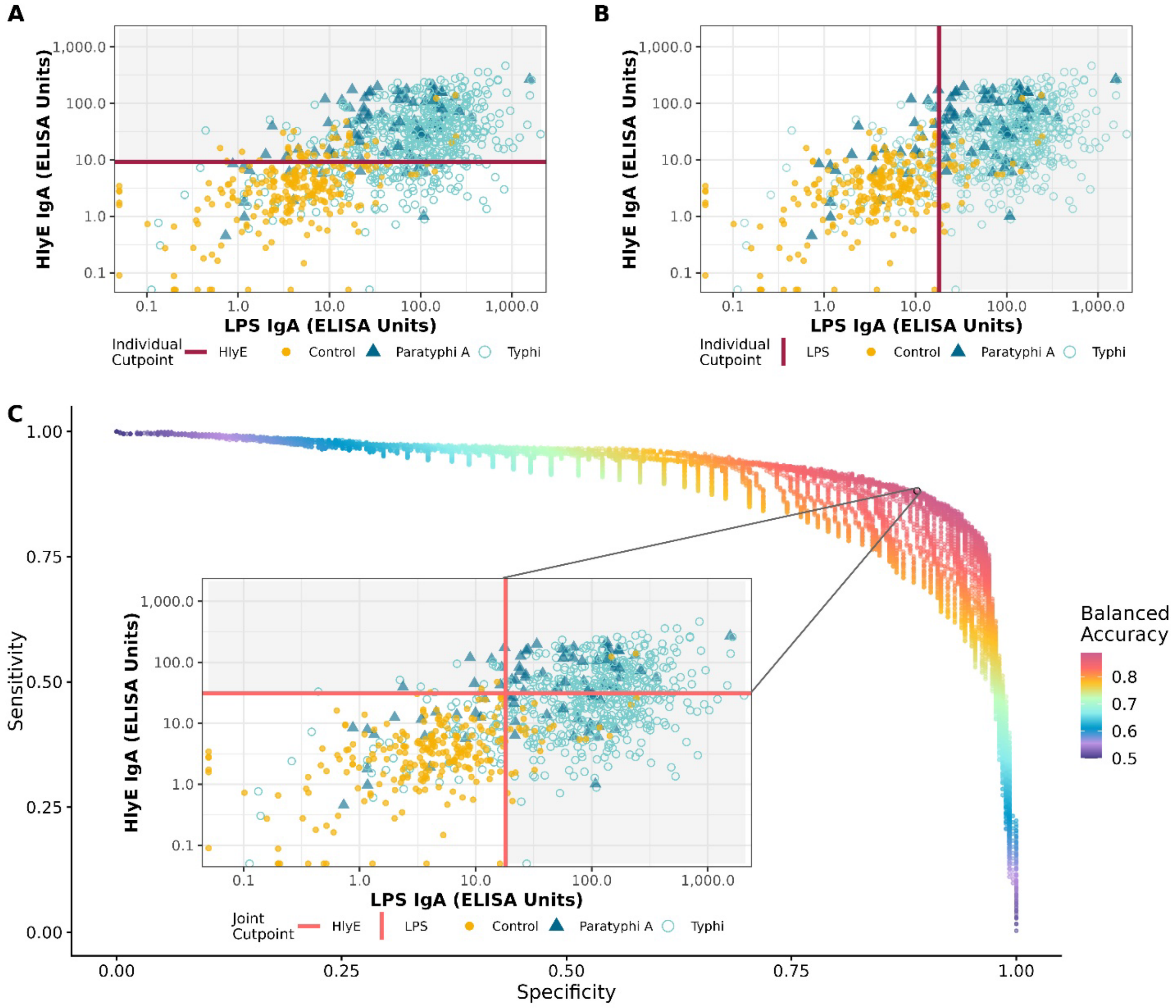
